# Prognosis of aneurysmal subarachnoid hemorrhage not altered with transatlantic airplane transfer. A bicentric matched case–control study

**DOI:** 10.1101/2024.02.06.24302429

**Authors:** Frédéric Martino, Milan Trainel, Jessica Guillaume, Aurélien Schaffar, Simon Escalard, Adrien Pons, Nicolas Engrand

## Abstract

**Background:** The treatment of a ruptured aneurysm, in a center with expertise in aneurysmal subarachnoid hemorrhage (aSAH), is recommended preferably within 24 to 72 hr. We assessed the impact of long-distance aeromedical evacuation in patients presenting aSAH in a remote island without neuro-interventional capacities.

**Methods:** This was a case–control study of patients with aSAH flown from a French Caribbean island (Guadeloupe) to Paris, France (6750 km), for neuro-interventional and neuro-ICU management and identical patients from the Paris region over a 10-year period (2010 to 2019). The two populations were matched on age, sex, World Federation of Neurological Surgeons score, and Fisher score. The primary outcome was the 1-year modified Rankin Scale (mRS) score divided into two categories: good outcome (mRS 0 to 3) and poor outcome (mRS 4 to 6). A cost study was added.

**Results:** Among 128 consecutive aSAH transferred from Guadeloupe, 93 could be matched with 93 patients with aSAH from the Paris area. The median [Q1,Q3] time from diagnosis to securing the aneurysm was 48 hr [30,63] in the Guadeloupe group versus 23 [12,24] in the control group (p<0.001). The rate of good clinical outcome (1-year-mRS ≤ 3) was 75% in the Guadeloupe group and 82% in the control group (p=0.1). The groups did not differ in 1-year mortality (18% vs 14%, p=0.5) and duration of mechanical ventilation. However, Guadeloupe patients more frequently required mechanical ventilation (59% vs 38%, p<0.001) and external ventricular drainage (55% versus 39%, p=0.005) than the control group, although the number of hydrocephalus events did not differ. The additional cost of treating a Guadeloupe patient in mainland France was estimated at 7580 euros, or 17% of the estimated cost in Guadeloupe.

**Conclusions:** Long distance aeromedical evacuation of Guadeloupe patients with aSAH resulted in a 25-hr increase in median embolization time but had no effect on mortality or functional prognosis at 1 year.

## Introduction

Spontaneous subarachnoid hemorrhage (SAH) accounts is the most common cause of neurovascular morbi-mortality.^1,2^ Its overall incidence is estimated at 6/100,000 person-years in international studies^3^ and 3.3/100,000 person-years in Martinique, a French Caribbean Island.^4^ Its etiology is related to a ruptured aneurysm in 85% of cases,^1^ and the management of aneurysmal SAH (aSAH) relies primarily on endovascular coiling of the ruptured aneurysm.^5^ Although the time to secure the aneurysm is not fully standardized, authors recommend coiling before 72 hr^1,2,6^ because the incidence of aneurysm re-bleeding is highest within 24 to 72 hr of the initial hemorrhage^7–9^ and is accompanied by significantly increased morbi-mortality.^2,6,8^ Coiling techniques are recommended to be performed in expert centers (defined as institutions treating more than 35 patients per year) and the prognostic benefits of transferring patients from low-volume to high-volume centers is well established.^2,6,10^ Hence, all over the world, patients have to be transferred to a sometimes distant referral center, with all the attendant organizational challenges and risks.^11^ Since 2010, in Guadeloupe, a French West Indies island of 440,000 inhabitants, all patients with aSAH have been transferred to the Rothschild Foundation Hospital (RFH), a high-volume referral center in Paris, for securing the aneurysm (coiling or surgical clipping) and management in the neuro-intensive care unit (neuro-ICU) (15-30 patients per year). With an agreement between the University Hospital of Guadeloupe (UHG) and airlines, this transfer is organized promptly by a transatlantic commercial flight. Nevertheless, this airplane transfer causes a mandatory additional delay from the aneurysm rupture to its treatment. In addition, the physiologic changes associated with spending several hours in a cabin pressurized to 0.75 to 0.8 atmosphere (atm) and the complexity of intensive care in the airplane could increase morbidity in the patient already weakened by the aSAH. Therefore, the question is to weigh the benefit of inter-hospital networks and standardized medical transport procedures against the risk of complications induced by the delay and conditions of the air transport itself.

We performed a retrospective bicentric case–control study comparing patients with aSAH who were air-transferred from Guadeloupe to Paris and patients from the Paris region who were cared for in the same high-volume referral center. The 2 populations were matched on their characteristics and severity of the aSAH: age, sex, World Federation of Neurological Surgeons (WFNS) score, and Fisher score. Our primary objective was to quantify the degree of disability and dependence between the 2 groups by assessing the 1-year modified Rankin Scale (mRS) score divided into 2 categories: good outcome (mRS 0 to 3) and poor outcome (mRS 4 to 6).

The research was approved by the Rothschild Foundation Hospital review board (IRB 00012801, study no. CE_20221122_3_NED). The study was reported according to the Strengthening the Reporting of Observational Studies in Epidemiology (STROBE) guidelines (http://www.equator-network.org).

## Patients and Methods

### Patient management

Once aSAH was diagnosed, patients were cared for according to the local guidelines, which met current literature and international guidelines.^2,6,12^ Briefly, systemic hemodynamic parameters were monitored and controlled by antihypertensive or vasopressor drugs (systolic blood pressure < 160 mmHg), hydration and natremia were normalized, oxygenation was controlled by mechanical ventilation if necessary, cerebral perfusion was monitored by transcranial Doppler ultrasonography (TCD), enteral nimodipine was initiated (720 mg/day), and external ventricular drainage (EVD) was implemented in case of threatening hydrocephalus (diagnosed on CT-scan). Then, patients were air-transferred with a dedicated medical team on a commercial flight, departing each time in the evening and arriving in Paris the next morning (6750 km, 8-hr 40-min flight, 14 hr total travel time). Neuro-interventional management of the ruptured aneurysm was performed on the first day in Paris. CT-scan was systematically performed in the angio suite before starting the endovascular procedure, to check for potential complications during the transfer (rebleeding, hydrocephalus, external shunt mobilization), as an institutional standard of care. Only patients with compressive hematoma were not considered for embolization and underwent neurosurgical clipping.

Furthermore, embolization is never ruled out in severe SAH, except for patients who die before embolization. Subsequently, patients were hospitalized in the neuro-ICU for at least 1 week, and management was in line with international guidelines (including cerebrospinal fluid shunting, prevention and treatment of cerebral vasospasm, and infectious complications).^6,12^ Patients from the Paris region received the same management upon arrival at the RFH.

### Study design

#### Patient selection

We enrolled consecutive patients admitted for aSAH between January 1, 2010 and December 31, 2019 who met the following criteria: 1) aSAH confirmed by CT-scan or MRI, 2) age ≥ 18 years, and 3) admitted to the neuro-ICU of the RHF for embolization of the ruptured aneurysm. Exclusion criteria were 1) age <18 years, 2) non-aneurysmal SAH, and 3) decision to withhold or withdraw treatment before admission.

We enrolled as many patients as possible who were admitted to the UHG and transferred to the RHF (Guadeloupe group). Then, for matching (control group), we identified patients in the RHF database who were admitted during the same period and were from the Paris region. Guadeloupe and control patients were matched on 2 standard demographic criteria (sex and age) and the 2 most widely used SAH severity scores: clinical (WFNS score) and CT-scan (Fisher score). Each case was matched to one control. The age criterion was extended to ± 7 years because it allowed the best compromise between number of pairs and clinical relevance.

#### Outcome

The primary outcome was the proportion of patients with mild to moderate disability at 1 year after admission among survivors, defined by a 1-year mRS score 0 to 3. The 1-year mRS score was extracted from patient follow-up consultation letters from neuroradiologists. If this score was not available, the treating physician was contacted. Patients could not be contacted directly because of the non-interventional nature of the study. Because this was a retrospective study, the mRS collection range was extended to 1 year ± 2 months.

Secondary endpoints were 1) time to secure the aneurysm since diagnosis (time of diagnosis defined by the initial CT-scan or MRI), 2) complications related to air transport, 3) complications during management in the neuro-ICU, 4) duration of mechanical ventilation and 28-day ventilator-free days, and 5) neuro-ICU length of stay. A cost study was added to assess the additional cost of treating a Guadeloupe patient in mainland France.

#### Statistical analysis

The matching involved using the Hopcroft-Karp matching algorithm to define the maximum number of patient pairs such that a patient in one group could be matched with only one patient in the other group. Analyses involved using R v4.0.3 (available at http://www.r-project.org/). Descriptive analyses are presented as numbers (percentages) for categorical variables and mean (SD) or median [Q1-Q3] for quantitative variables. Missing data for each variable are presented. Comparison between 2 categorical variables involved a McNemar test, and more than 2 categorical groups the Cochran Q test. Comparison between a quantitative variable and a qualitative variable involved a paired Student *t* test (or paired Wilcoxon test, as appropriate). The effect of air transfer on 1-year-mRS score ≤ 3 was analyzed by conditional logistic regression, with or without adjustment for aneurysm securing time. Results are reported as adjusted odds ratios (OR) with their 95% confidence interval (95% CI). Two-tailed p < 0.05 was considered statistically significant. Because of the exploratory nature of the study, we did not adjust for multiple comparisons.

## Results

Over the study period, 131 adults were admitted to UHG with confirmed aSAH: 128 were transferred to the RHF, and 3 were not transferred because of decisions to withdraw life-sustaining treatments before admission (n=2) and 1 administrative problem (Fig. 1). Over the same period, 659 adults were admitted to the RHF with confirmed aSAH, including the 128 from the UHG. After matching, 93 pairs were formed.

**Figure 1:**
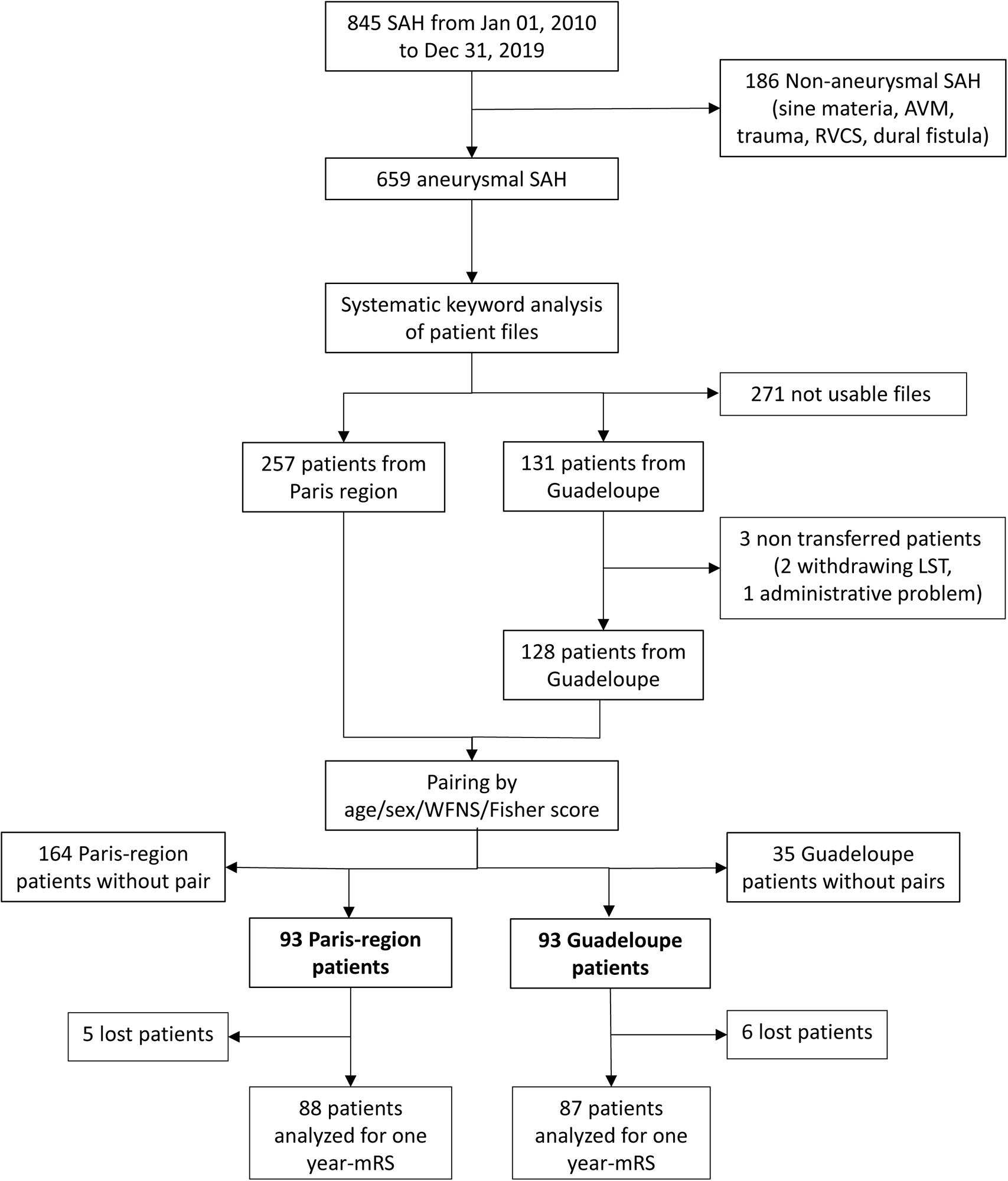
Flow chart. SAH: subarachnoid hemorrhage; AVM: arterio-venous malformation; RVCS: reversible vasoconstriction syndrome; WFNS: World Federation of Neurologic Surgeons; LST: life-sustaining therapies

### Patient characteristics

Of the 186 patients included, 120 (64.5%) were women, the mean age was 54 (SD 12) years, median WFNS score was 1 [1-3], and median Fisher score was 4 [3-4]. For the paired groups, the distribution of WFNS and Fisher scores was exactly the same for the two groups (Table 1 and SM1). Overall, 88% of ruptured aneurysms were located in the carotid system, many on the anterior communicating artery (SM 2). One patient in the Guadeloupe group did not benefit from aneurysm embolization (death before embolization) versus 4 patients in the control group (3 patients died before embolization, and 1 patient had a clip placed at the same time as surgical evacuation of an intracerebral hematoma).

**Table 1:**
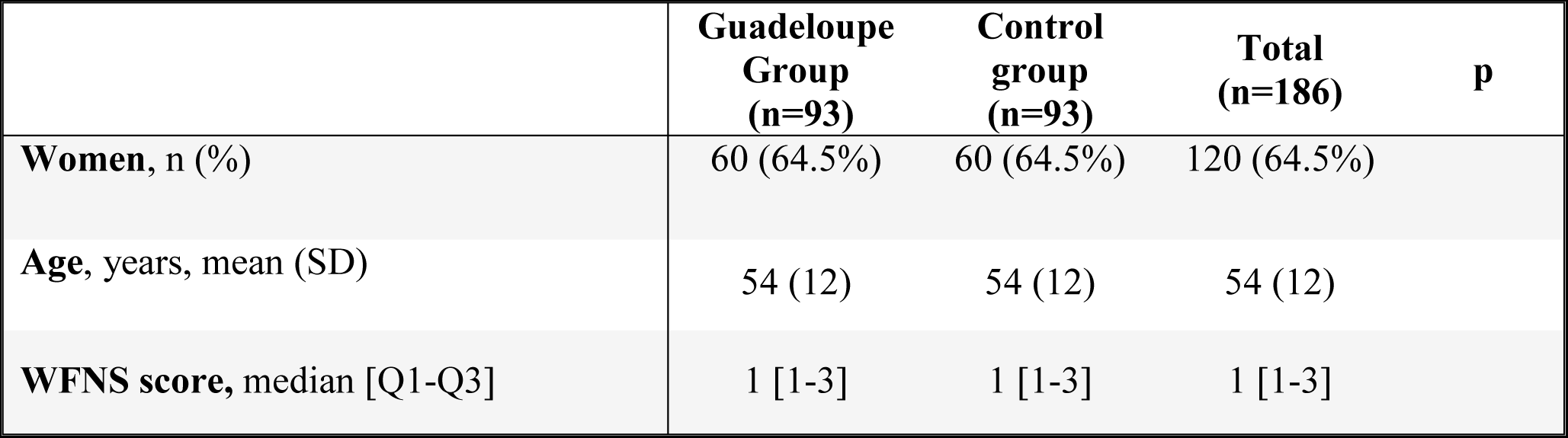
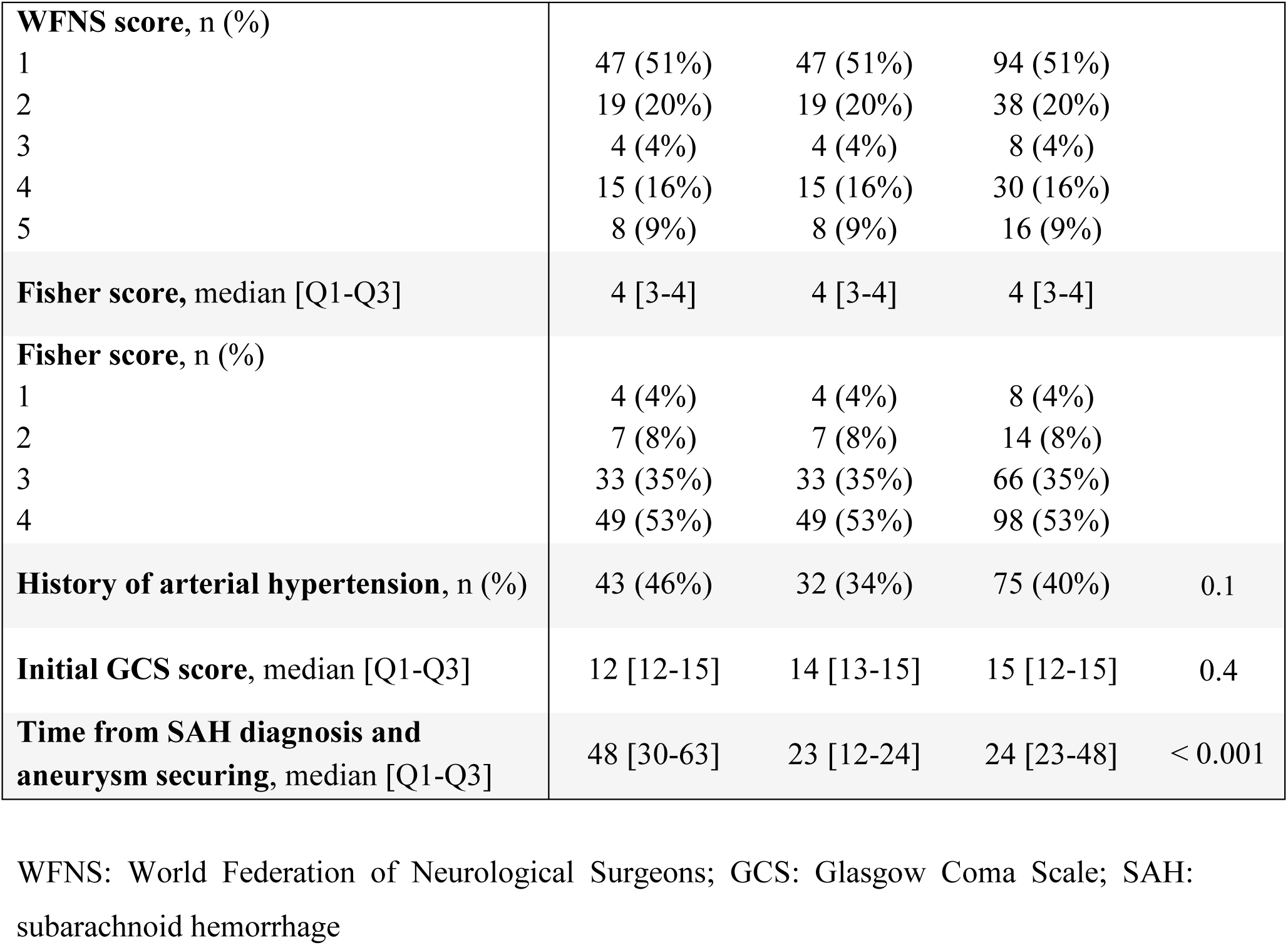
Characteristics of patients in the Guadeloupe and control groups.

### Primary outcome

Six patients in the Guadeloupe group and 5 patients in the control group were lost to follow-up, thus preventing evaluation of the 1-year mRS score. The Guadeloupe and control patients did not differ in proportion with 1-year mRS score ≤ 3: 75% (n = 65) versus 82% (n=72) (p = 0.1) (Fig. 2 and SM 3). The median 1-year mRS score was 1 [0-4] in the Guadeloupe group versus 1 [0-2] in the control group (p=0.08). The OR for a 1-year mRS score > 3 in the Guadeloupe group versus the control group was 2.14 (95% CI 0.87-5.26, p=0.1). After adjustment for time from diagnosis to embolization, the OR was 2.81 (95% CI 0.76-10.45, p=0.1).

**Figure 2:**
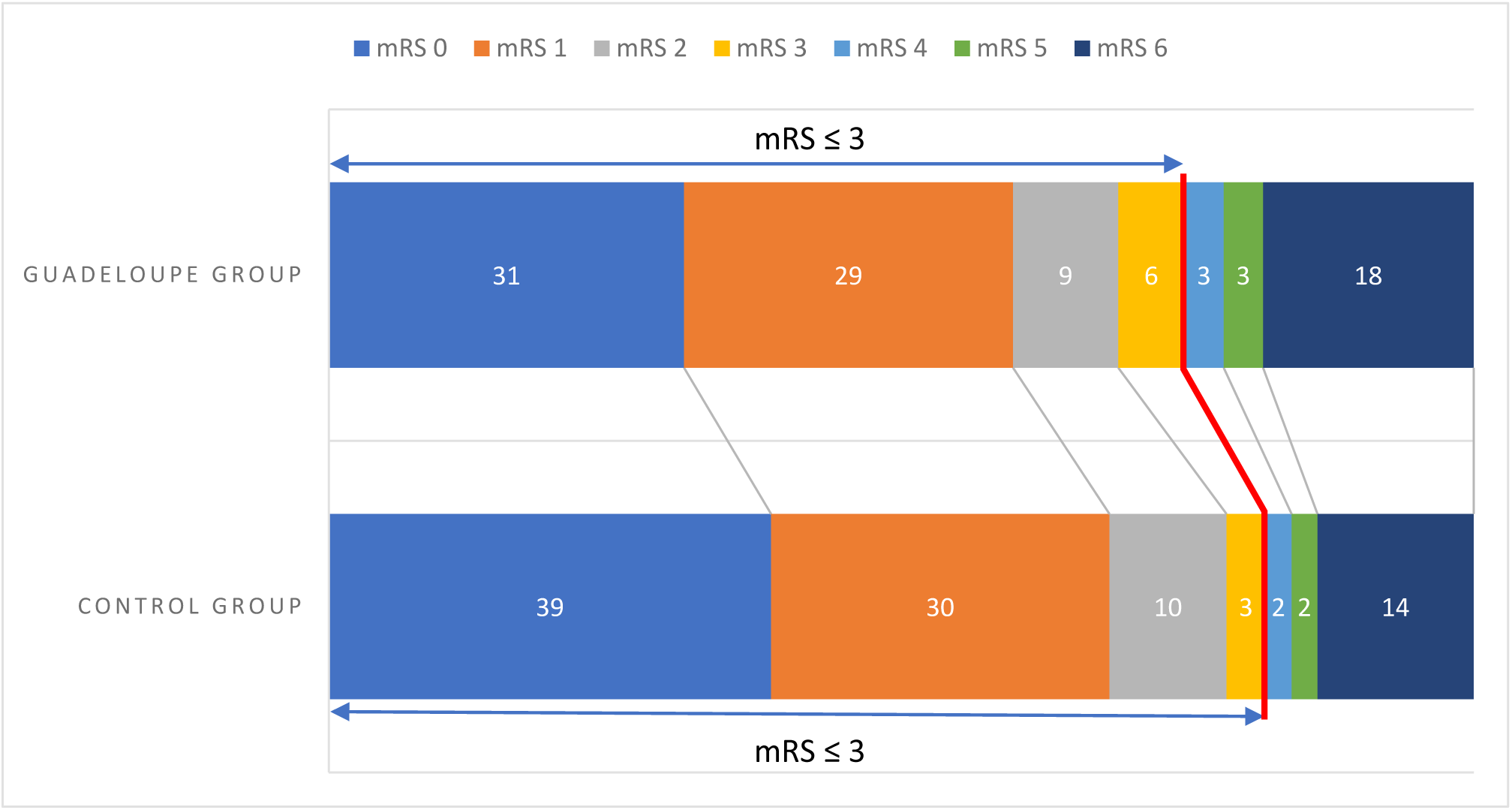
One-year modified Rankin Scale (mRS) score for the Guadeloupe and control groups (% of total) SM 1: Distribution of (A) World Federation of Neurological Surgeons (WFNS) and (B) Fisher scores for Guadeloupe and control groups SM 2. Location of ruptured aneurysms for all patients. NA: not applicable SM 3: One-year modified Rankin Scale (mRS) score for Guadeloupe and control groups MD: missing data The odds ratio for a 1-year mRS score >3 in the Guadeloupe group versus the control group was 2.14 (95% CI 0.87-5.26, p = 0.1). After adjustment for time from diagnosis to embolization, the odds ratio was 2.81 (95% CI 0.76-10.45, p = 0.1)

### Secondary outcomes: complications during and after air transfer

The Guadeloupe and control groups also did not differ in proportion of patients with 1-year mRS ≤ 2: 69% (n=60) versus 78% (n=69) (p=0.6, missing data for 6 and 5 patients, respectively).

Transport complications and comparison of ICU complications between the groups are in Tables 2 and 3. Only 1 aneurysm re-bleed was observed during air transfer (concomitant clinical deterioration and CSF hemorrhage in EVD, confirmed on arrival by CT-scan). Overall, 51 patients (55%) in the Guadeloupe group were intubated before the air transfer, 2 were intubated during the transfer and 2 were intubated after arrival in Paris. In the control group, 38% of patients were intubated during their stay in the neuro-ICU (p<0.001).

**Table 2:** Complications related to air transfer (Guadeloupe group only) (n=93)

|  |  |
| --- | --- |
| **Rebleeding, n (%)** | 1 (1%) |
| MD | 1 |
| **Epilepsy, n (%)** | 0 (0%) |
| MD | 2 |
| **Worsening GCS score, n (%)** | 2 (2%) |
| MD | 1 |
| **New neurological deficit, n (%)** | 0 (0%) |
| MD | 2 |
| **Initiation of noradrenaline therapy or dose increase ≥ 0.5 mg/hr, n (%)** | 9 (10%) |
| MD | 2 |
| **Orotracheal intubation, n (%)** | 2 (2%) |
| MD | 2 |
| **Increase in FiO<sub>2</sub> ≥ 25%, n (%)** | 1 (1%) |
| MD | 3 |

MD: missing data; GCS: Glasgow Coma Scale, EVD: external ventricular drainage

**Table 3:**
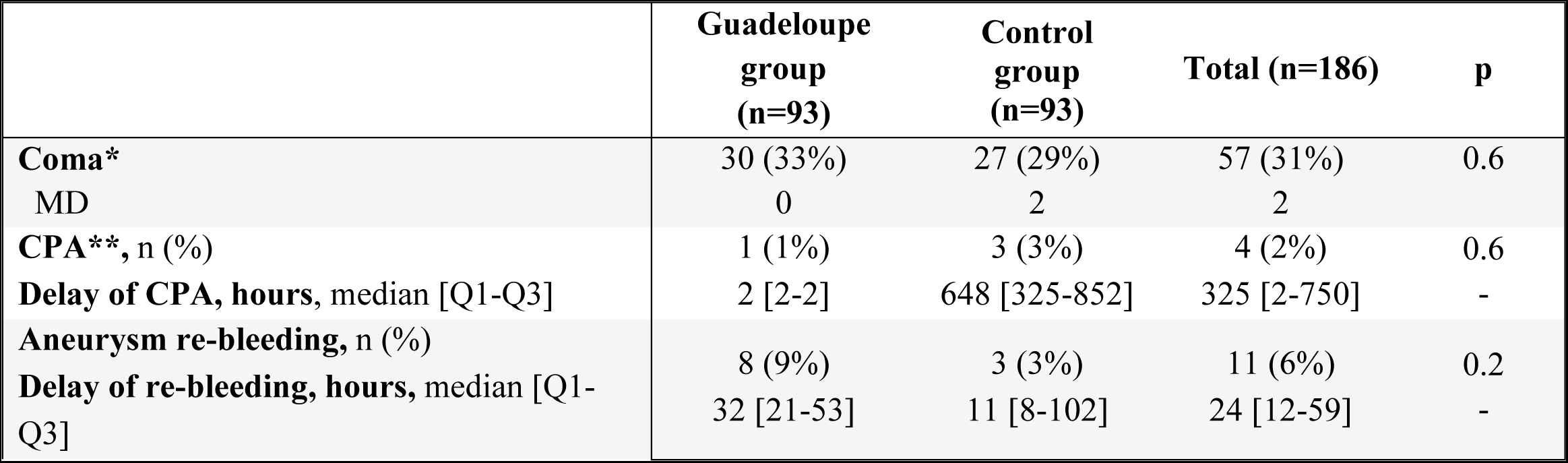
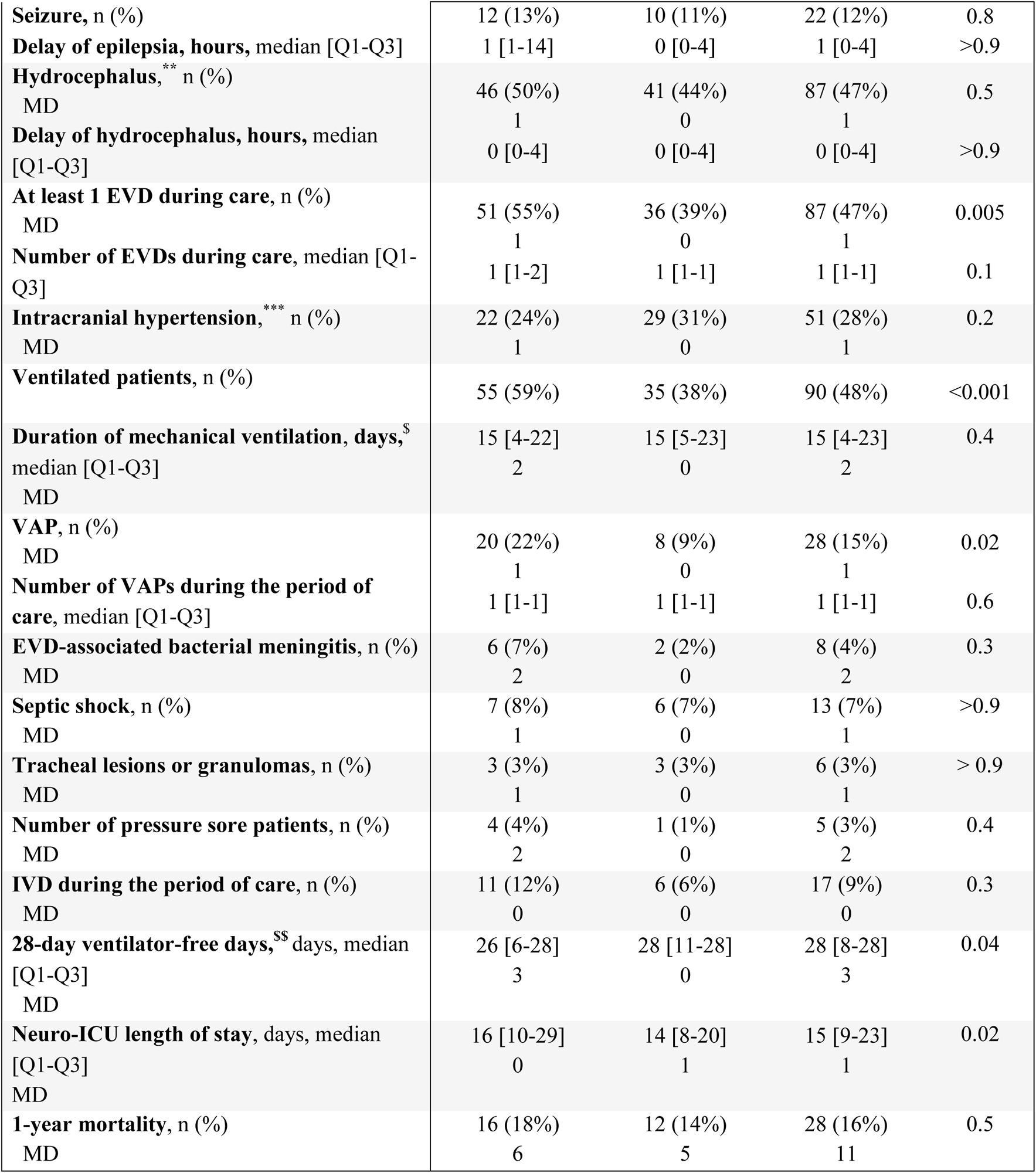
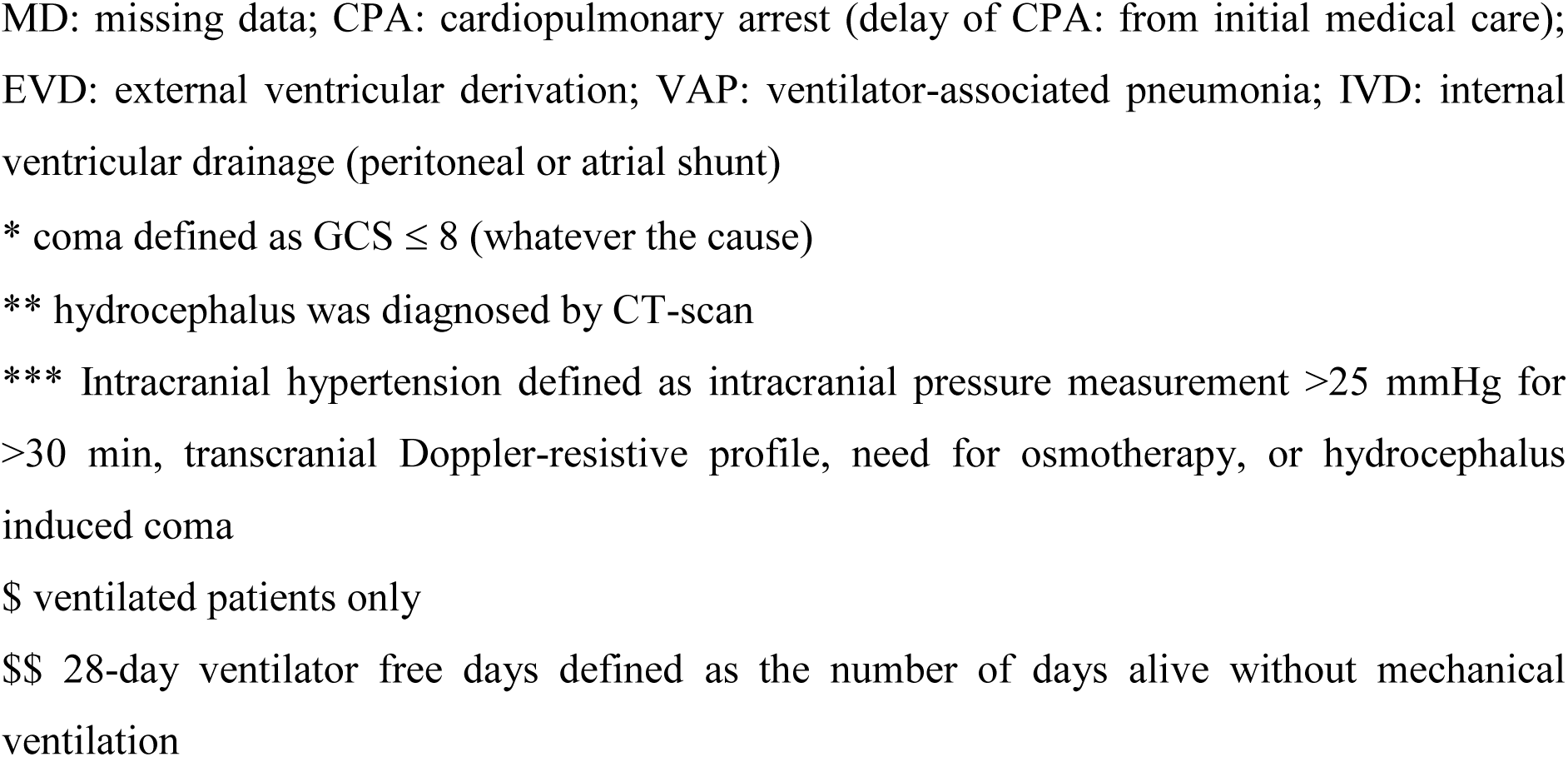
Comparison of intensive care unit (ICU) complications between groups.

The median time from diagnosis to aneurysm embolization was significantly longer for Guadeloupe than control patients: 48 hr [30,63] versus 23 hr [12,24] (p<0.001).

A total of 51 patients (55%) from Guadeloupe had at least one EVD during their treatment course as compared with 36 patients (39%) in the control group (p=0.005), while the number of hydrocephalus events did not differ between the Guadeloupe and control groups (46 [50%] and 41 [44%] patients, p=0.5) (Table 3). No EVD was replaced at the RFH when the patient arrived from Guadeloupe (due to ventricular dilatation or EVD mobilization or obstruction during the flight).

The number of patients with at least one episode of ventilator-associated pneumonia (VAP), and the ICU length of stay were significantly higher for Guadeloupe than control patients, and the 28-day ventilator-free days was lower (Table 3). However, the groups did not differ in duration of mechanical ventilation or 1-year mortality (18% vs 14%).

### Cost study (Table 4)

Cost data were available for 62 pairs of patients. In this subset, the total cost of hospital stay was 50768 versus 28461 euros for Guadeloupe and control groups (no statistical comparison possible), and the mean lengths of stay in the neuro-ICU differed (17 vs 14 days, p=0.05). If all care for Guadeloupean patients had been provided locally, for the same length of hospital stay, and taking into account that healthcare costs are 20% higher in Guadeloupe than in Paris, the extrapolation of costs in Guadeloupe was 43188 euros/patient, for an estimated additional cost of 7580 euros/patient, or 17% of the estimated cost in Guadeloupe.

**Table 4:**
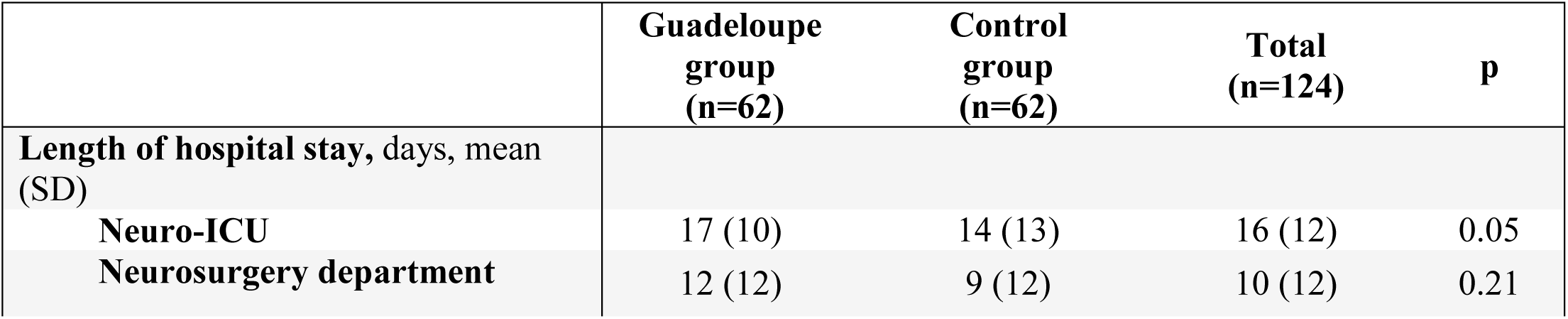
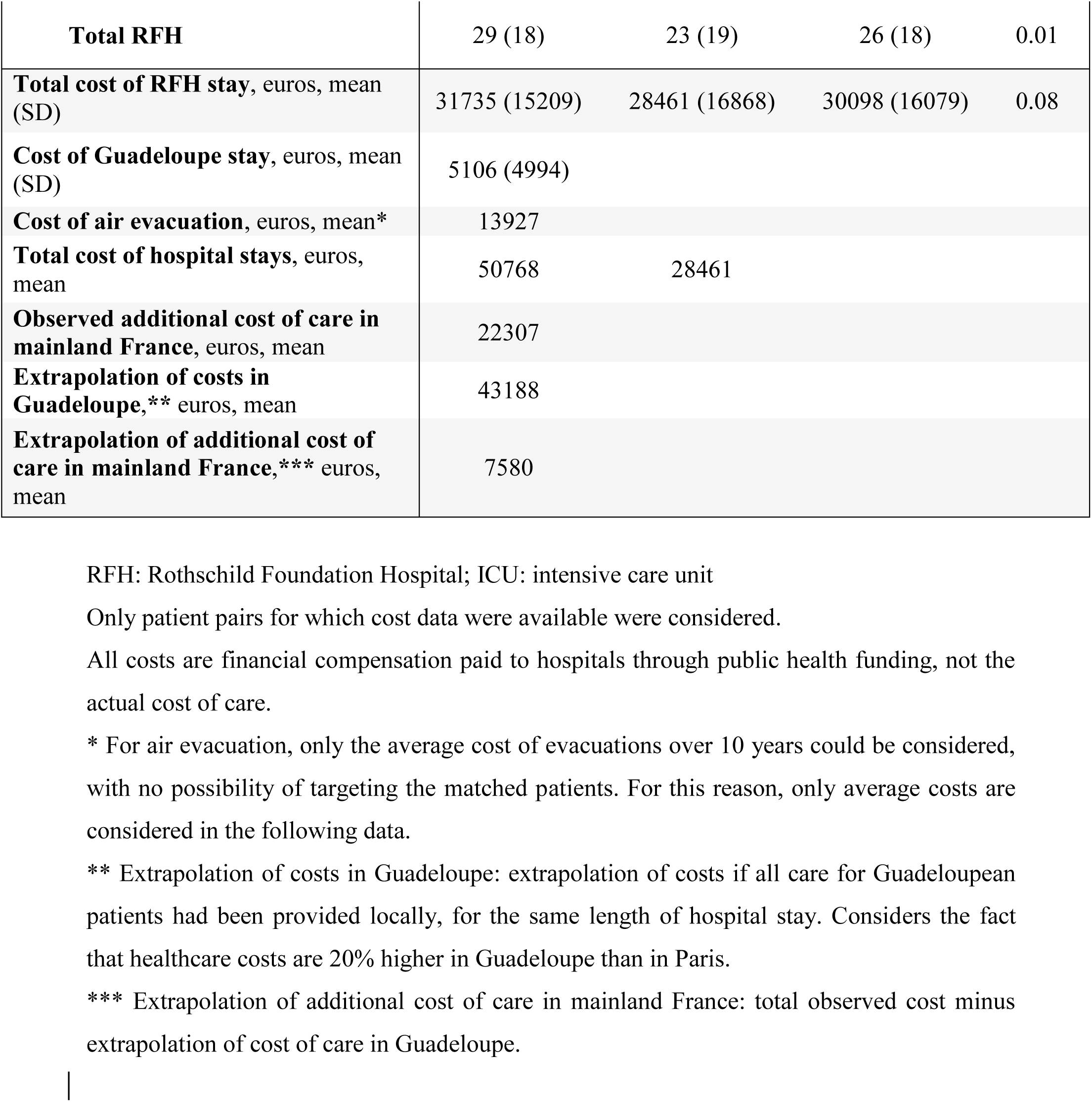
Cost study.

## Discussion

We found no difference in 1-year functional outcome and mortality between patients with aSAH who were transferred by airplane from Guadeloupe to Paris and matched aSAH patients from the Paris region, including after adjustment for embolization delay. However, patients from Guadeloupe had significantly longer delay to securing the aneurysm as well as higher frequency of mechanical ventilation and ventricular drainage than control patients, despite identical disease severity and similar proportions of hydrocephalus.

Mejdoubi et al. studied a population of 119 aHSA patients from Martinique (another Caribbean island very close to Guadeloupe) over 10 years, 91 of whom were air-transferred to mainland France (also to the RFH), with the alternative of surgical treatment of the aneurysm in Martinique or conservative treatment for patients with the most severe disease.^13^ The proportion of patients with mRS score ≤ 3 and mortality at hospital discharge was 80% and 9%, respectively, for patients transferred to metropolitan France as compared with 75% and 18% at 1 year in our series. This difference might be explained by the timing of the result measurement and by patients with the most severe disease being not transferred to mainland France.

Weyhenmeyer et al. showed no difference in mortality or functional prognosis between transferred and non-transferred aSAH patients.^11^ Air transport was associated with worse functional prognosis than ground transport, except for patients with the most severe disease. However, the distances flown were significantly shorter than in our study (mean 120 [SD 80] km for the air-ambulance group). With its comparative design, our paired-group study confirms that the need for transatlantic air transfer did not entail a loss of chance in the management of aSAH in a Caribbean patient, all the more so because our patients were not selected for inclusion on the basis of clinical or CT-scan disease severity. Also, when combining the Mejdoubi et al. series with our own (194 total patients), only 1 re-bleed was observed during air transfer.

Conversely, other studies have shown that simply having to transfer a patient with SAH from a general to a tertiary-care center can lead to deterioration in functional prognosis and mortality.^10,14,15^ However, these studies are dated and the conditions of medical transport were not specified. In addition, early studies reported up to 20% re-bleeding during transfer,^16^ whereas we observed a lower overall rate of re-bleeding at the time of embolization: 8/93 and 3/93 patients in the Guadeloupe and control groups (p=0.2). This finding could be due to better control of hypertension and coagulopathies, as recommended by recent guidelines.^2,6,12^ In terms of the impact of delayed embolization caused by air evacuation, the prognosis may not differ between securing the aneurysm at day 0, 1, 2 or 3^17,18^ perhaps because of the 6- to 12-hr earliness of re-bleeding in most cases.^7,11^ Thus, only embolization within the first 12 or even 3 hr would allow for a real reduction in re-bleeding, ^7,19,20^ which was not achieved in our series, including in our control group.

Overall, the prognosis of patients in our series seemed as good as that recently published by other teams in developed countries, with a frequency of 70% to 88% mRS score ≤ 3 and 10% to 43% mortality.^5,21,22^

Beyond the pure safety aspect, the other objective of our study was to evaluate whether the context of aeromedical evacuation led to a different management than for patients directly admitted to the RFH. About 10% of Guadeloupe patients who underwent EVD had no established diagnosis of hydrocephalus, whereas in the metropolitan France group, about 12% of patients with hydrocephalus benefited from a wait-and-see attitude to avoid this invasive procedure. Implementing EVD aimed to ensure air-transport safety, sometimes “prudently”, to avoid in-flight intracranial hypertension, which could have marked effects. Similarly, the number of intubated patients was significantly higher in the Guadeloupe than control group, 89% of them before the flight. Actually, all patients who underwent EVD under general anesthesia just before medical transfer remained sedated after the procedure. This prevented accidental removal of the EVD in patients who could be confused and agitated during transport.

This use of prophylactic invasive therapies was not harmless because the number of VAP cases was significantly higher in the Guadeloupe than control group. However, the frequency of EVD-related meningitis did not differ between the groups, nor did the number of EVDs per patient among those undergoing at least one EVD. Regardless, the morbidity of EVD is well established.^23^

Air travel causes physiological changes that can lead to specific complications. To limit hypobaric hypoxia, the Federal Aviation Administration requires airlines to pressurize civil flight cabins to a level equivalent to an altitude < 8,000 feet^24^ (i.e. a pressure ≥ 0.75 atm), which offers a reasonable compromise between reducing the partial pressure of oxygen in ambient air and the economic cost of pressurization. In a healthy individual, this would be equivalent to breathing sea-level air at FiO2 15%,^24^ resulting in a 35 mmHg decrease in PaO2 to about 60 mmHg.^25^ These levels can become critical in a non-intubated patient at risk of low cerebral blood flow, as can occur after SAH (e.g., in situations of intracranial hypertension or cerebral vasospasm). In a swine model of traumatic brain injury, a simulated 4-hr aeromedical transfer at 8,000 ft induced a significant decrease in mean arterial pressure, brain tissue O_2_ pressure, and regional cerebral blood flow.^26^ Moreover, the autonomic neurological system could be disturbed, leading to a greater variation in blood pressure than under normobaric conditions,^27^ which may explain why an increase in catecholamine doses during flight was necessary in 10% of our patients. However, a study of head injury patients showed that aeromedical evacuation was ultimately beneficial, with the benefits in terms of prompt care outweighing the risks of flying, with an even greater benefit for patients with the most severe disease.^28^ On the other hand, a decrease in ambient pressure could theoretically lead to a decrease in the aneurysm’s transmural pressure, thus increasing the risk of re-bleeding, but this phenomenon has never been proven.

In the subset of patients included in the cost study, as compared with control patients, Guadeloupe patients stayed longer in the neuro-ICU but not in the neurosurgery ward, perhaps because of the excessive need for weaning from mechanical ventilation and EVD. The identical duration of mechanical ventilation and IVD placements in both groups, with higher number of mechanical ventilation and EVD procedures in the Guadeloupe than control group (Table 3), support this hypothesis. In addition, some Guadeloupe patients may have stayed longer in Paris because of the constraints associated with organizing their return to Guadeloupe, which is less codified than emergency air evacuation. In any case, the additional cost of care, estimated at 17% of Guadeloupe’s costs, seems acceptable in view of the service provided to the patient.

The strengths of our study are that our primary endpoint (1-year mRS score) was the gold standard of neurovascular studies^5,29^ and that comparison of matched series represents the most scientifically valid means of assessing the effect of medical air evacuation on patient prognosis. Indeed, a randomized prospective study in this context would not be possible, even if the embolization procedure were available in Guadeloupe. Moreover, we studied the additional cost of air evacuation and placed it into perspective with the total cost of the initial care of our patients.

However, our study has some limitations. First, the selection of eligible patients was difficult because of the retrospective nature of the study. We needed to develop an algorithm to scan reports for keywords and to correct discordant WFNS and Fisher scores before matching patients. In addition, we could not determine the exact time in hours between aneurysm rupture and diagnosis. Nevertheless, the time between diagnosis and embolization was reliable. Second, pairing necessarily involved some degree of patient selection, with risk of selection bias (in particular, exclusion of patients with the most severe disease because of lack of air transfer). Nevertheless, this risk was limited by the very few patients who were not evacuated or embolized because of their disease severity and that matching was based only on the possibility of matching criteria and never on disease severity. Finally, the distribution of WFNS and Fisher scores was similar to that found in other aSAH studies, as was the distribution of anterior and posterior circulation aneurysms.^5,21^ Third, we did not record the prevalence of diabetes, body mass index or occurrence of cerebral vasospasm.

## Conclusions

This study showed no difference in the long-term functional prognosis of patients with aSAH in Guadeloupe and those from mainland France who underwent treatment directly at the referral center, despite aeromedical evacuation and a 25-hr longer median embolization time for the former. Because of its robustness, this study reinforces the view that the need for long distance air-evacuation of Guadeloupean patients does not entail a loss of chance for the patient, at an acceptable additional cost. These results may be transferable to other geographical situations worldwide, provided that the process is sufficiently well prepared in terms of flight availability, aircraft organization, mobilization of accompanying medical teams and patient conditioning (EVD, ventilatory assistance, etc.).

## List of abbreviations

aSAH: aneurysmal subarachnoid hemorrhage
ICU: intensive care unit
mRS: modified Rankin scale
OR: odds ratios
RFH: Rothschild Foundation Hospital
UHG: University Hospital of Guadeloupe
WFNS: World Federation of Neurologic Surgeons
CI: confidence interval

## Declarations

The research was approved by the Rothschild Foundation Hospital review board (IRB 00012801, study no. CE_20221122_3_NED).

## Consent for publication

Not applicable.

## Availability of data and material

The datasets used and/or analyzed during the current study are available from the corresponding author on reasonable request.

## Authors’ contributions

FM, MT, AS, AP and NE contributed to study design, data collection, data acquisition.

FM, MT, JG, AS, SE, AP and NE contributed to data analysis, interpretation and drafting the manuscript.

All authors read and approved the final manuscript.

## Data Availability

Data available on request within reasonable limits

## Acknowledgements

Laura Smales (proofreading), Xavier Marcatel and Lucile Sénicourt (data scientists, RFH), Dr Djamel Bouamama (Medical Informatics Department, UHG), Raphael Juraver and Prof P. Portecop (Emergency medical service, UHG), Dr Jean-David Pommier (ICU, UHG).

